# Characterization of the Urine Proteome Using the Olink Explore 3072 Platform

**DOI:** 10.64898/2025.12.08.25341840

**Authors:** Marisa Meier, Mona Schoberth, Yong Li, Oleg Borisov, Burulca Uluvar, Christopher Willeke, Heike Meiselbach, Kai-Uwe Eckardt, Oliver Schilling, Anna Köttgen, Stefan Haug

## Abstract

Urinary proteins are valuable biomarkers for physiological processes and early detection and monitoring of systemic and kidney diseases. However, comprehensive quantification and characterization of the urine proteome using high-throughput platforms remains limited.

Using an affinity-based assay for large-scale proteomics, the Olink Explore 3072 platform, we quantified 2,868 proteins in urine samples from 1,268 participants of the German Chronic Kidney Disease (GCKD) study.

Protein detectability in urine, defined as the percentage of samples with measurements above the limit of detection, showed a bimodal distribution: 659 (23%) proteins had high (>80%) and 1,336 proteins (47%) had low (<20%) detectability. Over 95% of highly detectable proteins were confirmed as present in urine by mass spectrometry. They were enriched for extracellular, exosomal, and cell-surface localization, and showed tissue-specific expression in hepatocytes, immune cells, and proximal tubule cells.

Significant associations of protein levels with sex (921 proteins), BMI (367 proteins) and age (157 proteins) showed plausible relationships such as prostate-specific antigen and male sex. Unsupervised correlation analysis of protein levels detected multiple biologically meaningful protein clusters, reflecting shared cellular compartments (e.g., lysosomal origin), tissue of origin (e.g., liver), and molecular interactions (e.g., receptor-ligand or chaperone-transporter complexes).

In conclusion, the Olink Explore 3072 platform is well-suited for large-scale urine proteomics and identifies biologically plausible representations of ongoing processes in health and disease. Our comprehensive characterization provides a valuable resource for biomarker discovery and future targeted studies of urine proteins.

**Translational Statement:** Urinary proteins reflect kidney-specific and systemic processes, with the potential to serve as disease biomarkers. We demonstrate that the Olink Proximity Extension Assay enables high-throughput urine proteome screening with sufficient specificity to identify biologically plausible protein signatures and demographic associations. By providing comprehensive data on protein detectability, variability, and clinical associations, our study serves as a reference resource for designing targeted proteomic investigations, facilitating the development of urine-based biomarker panels for different purposes, potentially including early detection of impaired kidney function, monitoring kidney disease progression, and predicting treatment response.

## Introduction

Urine contains proteins from the kidneys, urinary tract, and systemic circulation^1^ and their composition changes with physiological processes, such as aging, and in states of disease, making them valuable biomarkers for kidney and systemic diseases such as CKD^2^ and cancer^3,4^. The non-invasive nature of urine collection enhances its potential for biomarker discovery and clinical testing^5^.

Over the past two decades, proteomic assay technologies have undergone significant improvements in coverage and throughput. In particular, affinity-based methods that utilize aptamers or antibodies binding to specific target proteins are now sufficiently time- and cost-effective to conduct large-scale population-based proteomic studies in plasma or serum^6,7^. Prominent examples using Olink”s Proximity Extension Assay (PEA) technology include the UK Biobank Pharma Proteomics Project^8^ (nearly 3,000 proteins in over 50,000 plasma samples) and the Human Blood Atlas^9^ (up to 5,400 proteins in over 8,000 samples). In urine, previous studies have used Olink”s technology with 96-protein panels^10–14^, or with high-coverage libraries but in small study samples (n=47-174)^15–17^.

In this study, we evaluate the feasibility and utility of detecting the complete Olink Explore 3072 library in a large set of urine samples from 1,268 individuals, providing comprehensive information on the detectability and variability of each protein. Detectability values were compared to mass spectrometry data, and highly detectable proteins were characterized in terms of their potential cellular origin and biological functions. Additionally, we examined associations between all detected proteins and demographic factors such as age, sex, and BMI, as well as patterns of protein co-regulation that are apparent in urine and represent a readout of ongoing physiological processes.

## Methods

### Urine samples, proteomic measurement, processing and quality control

We analyzed urine samples from 1,268 participants of the German Chronic Kidney Disease (GCKD) study^18^ with baseline estimated glomerular filtration rate (eGFR) 45-60 ml/min/1.73m^2^ and urine albumin-creatinine ratio (UACR) <300 mg/g. The study was approved by local ethics committees and all participants provided written informed consent.

Spot urine samples were centrifuged (1500g, 10 min) within 30 minutes of collection, then frozen at - 80°C and shipped to a central biobank. Proteomic profiling was performed using the Olink Explore 3072 Proximity Extension Assay (PEA) technology^19^ in two independent batches (n=769 and n=499), measuring 2,943 analytes targeting 2,926 unique proteins (**Figure 1A**). Following quality control (58 proteins with 100% QC warnings, five outlier samples and one sample with excessive warnings removed), 2,886 analytes representing 2,868 distinct proteins in 1,268 samples remained for analysis. Protein levels were corrected for urine dilution using the probabilistic quotient method^20^. Detectability values represent the percentage of urine samples in which a given protein was measured above its limit of detection.

**Figure 1:**
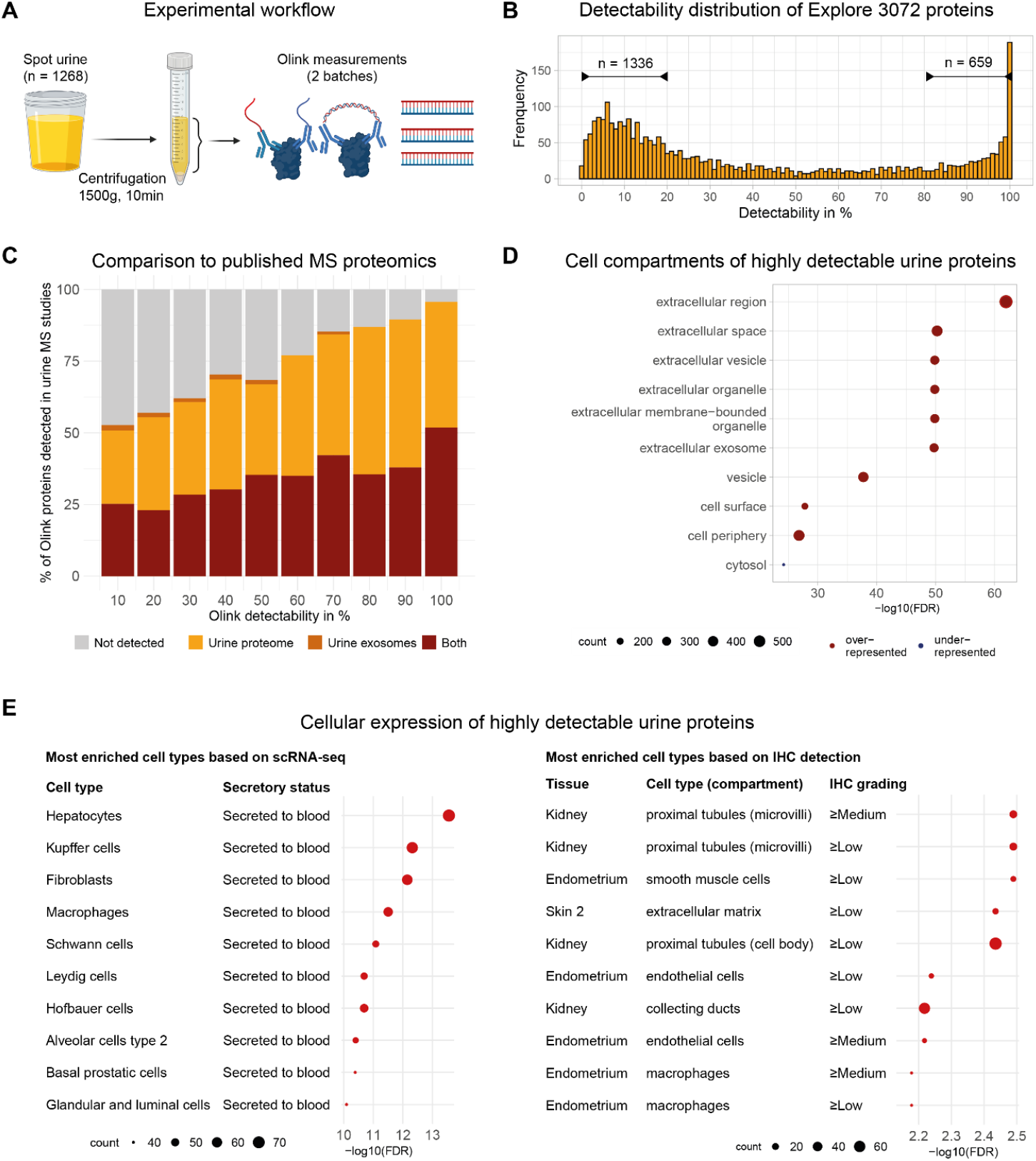
Detectability and characterization of the urine proteome using Olink Explore 3072. **(A)** Experimental workflow. Spot urine samples (n=1,268) were centrifuged and analyzed in two batches using the Olink Explore 3072 platform. **(B)** Distribution of protein detectability across both batches. **(C)** Percentage of proteins previously detected in mass spectrometry (MS)-based urine proteomics studies (overall urine proteome, exosomal fraction, or both), stratified by detectability categories. **(D)** GO-Term Enrichment of cellular compartments among highly detectable proteins (detectability >80%). **(E)** Enrichment analysis for highly detectable proteins. Left panel: enrichment for cell types and secretory status based on RNA expression profiles and HPA secretome data. Right panel: enrichment for tissue-specific cell types and cellular compartments based on immunohistochemistry data.

### Enrichment analyses

Highly detectable proteins (detectability >80%) were tested for enrichment of Gene Ontology (GO) terms, cell types with corresponding information about proteins” secretory status, and tissue-specific expression patterns based on immunohistochemistry data from the Human Protein Atlas^21^. Enrichment was assessed using Fisher”s exact test with Benjamini-Hochberg correction for multiple testing.

### Association with demographic characteristics

For associations of dilution-corrected protein levels with age, sex, and BMI we used multiple linear regression with all three variables fitted in the same model along with baseline ln(eGFR) in ml/min/1.73m^2^, ln(UACR) in mg/g and measurement batch as additional covariates. Only proteins with a false discovery rate (FDR) < 0.05 for one of the three tested demographic characteristics are reported.

### Correlation analyses

Protein levels were adjusted for potential technical and biological confounders by extracting standardized residuals from linear models including dilution correction factor, ln(eGFR), ln(UACR), age, sex, and batch. Pearson correlations were calculated for each pair of proteins, and proteins with at least one correlation r > 0.7 (n=217) were selected for unsupervised hierarchical clustering. Regularized partial correlations were estimated using graphical LASSO with EBIC model selection^22^ to identify potential direct protein-protein relationships and reduce false-positive correlations.

The full methods are provided in supplementary material.

## Results

Analyses of protein levels in our study sample of 1,268 GCKD participants indicated no significant differences in eGFR, UACR, or other relevant clinical characteristics across the two measurement batches (**Supplementary Table 1**). Proteomic analysis captured 2,868 distinct proteins measured by 2,886 analytes (**Supplementary Table 2)**.

### Hundreds of proteins show high detectability in urine

Detectability values for all proteins (**Supplementary Table 3**) were highly correlated between measurement batches (Spearman”s rho = 0.86). Combined detectabilities showed a bimodal distribution: 659 proteins (23%) with detectability >80% and 1,336 proteins (47%) with detectability <20% (**Figure 1B**). The eight Olink protein sub-panels showed substantially different detectability distributions (**Supplementary Figure 1**), with the cardiometabolic I panel containing the most highly detectable proteins (84% median detectability) and neurology II the least (12% median detectability). We observed that protein targets assayed across multiple panels strongly correlate, particularly for highly detectable proteins like C-X-C Motif Chemokine Ligand 8 (CXCL8, **Supplementary Figure 2**). Analyses stratified by median eGFR and UACR revealed negligible mean differences in detectability across proteins (0.17% for eGFR and 2.74% for UACR), and highly consistent detectability patterns with correlations of >0.99 for both (data not shown).

**Figure 2:**
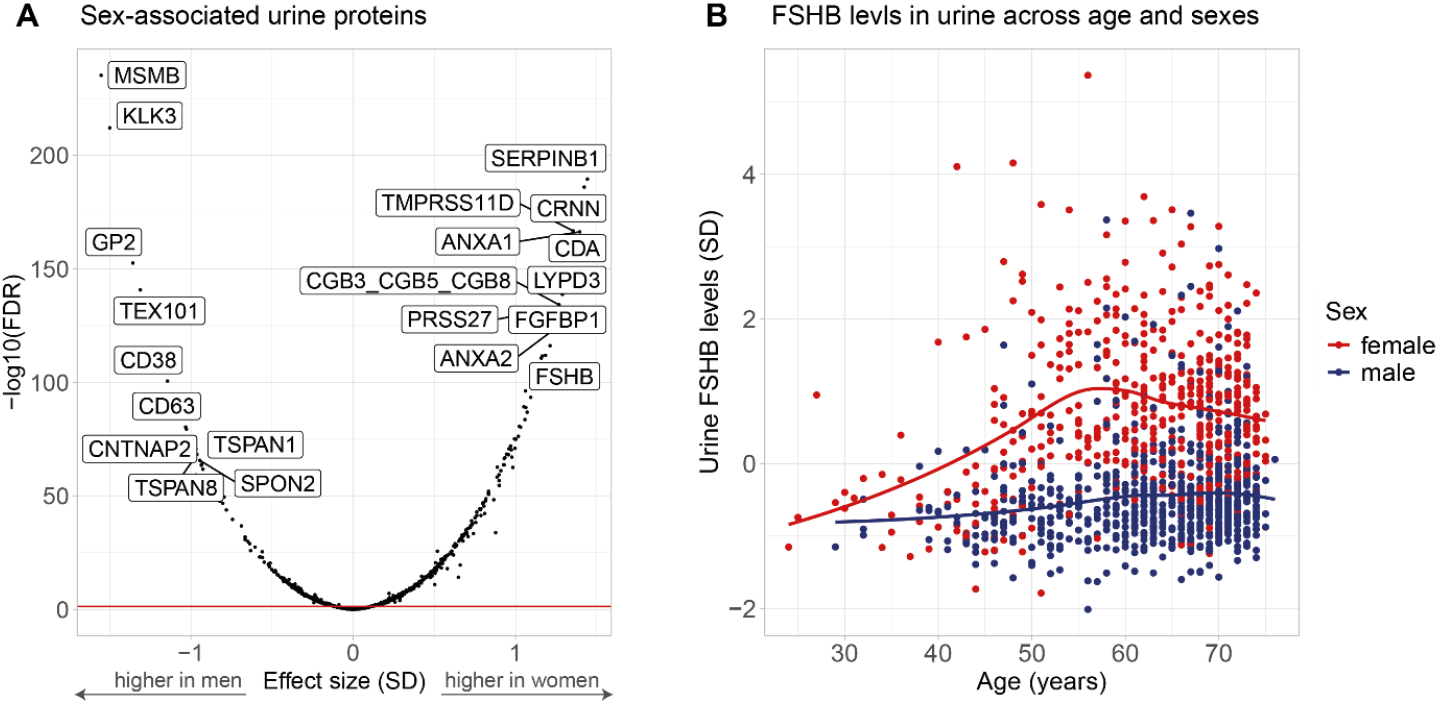
Association of urine proteins with demographic characteristics. **(A)** Association of dilution-corrected urine protein levels with sex from multiple linear regression models adjusted for covariates. The horizontal red line indicates the significance threshold (FDR = 0.05). Effect sizes are shown as standardized coefficients (SD, standard deviation units). **(B)** Dilution-corrected urine Follicle-Stimulating Hormone Subunit Beta (FSHB) levels stratified by age and sex (n=1,248). Curves represent LOESS-smoothed trends for men (blue) and women (red).

Over 95% (484/506) of proteins with detectability >90% were previously detected by mass spectrometry^1^, declining to 53% (391/742) for proteins with detectability 0-10% (**Figure 1C, Supplementary Table 4**). Highly detectable proteins were enriched for extracellular localization (79% extracellular, 48% exosomal, 30% cell surface), while intracellular proteins were underrepresented (**Figure 1D, Supplementary Table 5**). Functional enrichment analysis revealed overrepresentation of proteins with signal receptor activity (21%), as well as proteins involved in cell adhesion (34%) and immune system processes (40%) (**Supplementary Figure 3**). Cell type enrichment analysis identified macrophages, hepatocytes, and proximal tubule cells as potential main contributors to the highly detectable urine proteome (**Figure 1E**).

The variability of all analytes across urine samples was quantified with interquartile ranges (IQR; **Supplementary Table 6**). The most variable proteins included sex-specific proteins (e.g., Microseminoprotein Beta (MSMB), Prostate-Specific Antigen (KLK3)), proteins affected by common genetic variants (Prostate Stem Cell Antigen (PSCA), Histo-blood Group ABO System Transferase (ABO)), and many proteins involved in the immune response to external pathogens (**Supplementary Table 7, Supplementary Figure 4**).

### Various proteomic links to demographic factors

We examined associations (FDR < 0.05) between protein levels and demographic factors (**Supplementary Table 8**), while controlling for participants” eGFR and UACR. Significant protein associations with sex (n=921, **Figure 2A**) were more frequent than associations with BMI (n=367, **Supplementary Figure 5A**) and age (n=157, **Supplementary Figure 5B**). The leading sex-associated proteins included KLK3 and MSMB for men. For women, they included Leukocyte Elastase Inhibitor (SERPINB1), Cornulin (CRNN), and Follicle-Stimulating Hormone Subunit Beta (FSHB). KLK3 and MSMB are highly expressed in the prostate gland, while SERPINB1 and CRNN are expressed in leukocytes and vaginal epithelium, respectively. For FSHB, the beta subunit of the pituitary hormone FSH, our measurements clearly showed an increase in urine levels among women during perimenopause (**Figure 2B**).

### Protein correlation network highlights protein sources along the urinary tract

We selected 217 proteins with at least one high pairwise correlation (r>0.7) with another protein for network analysis (**Supplementary Table 9, Figure 3A**) and identified high-confidence protein relationships (**Supplementary Figure 6, Figure 3B**). The resulting network revealed multiple mechanisms of protein co-regulation: subnetworks of proteins aligned with shared expression in cell types found along the urinary tract, e.g. in sperm cells (ACRV1, LYZL2, SPINT3 and PAEP), urothelial cells (SPINK1, CCN5), immune cells (BTN3A2, ICOSLG, GDF15, OLR1) and squamous epithelial cells (CDA, SERPINB1, LYPD3, DSG3, KLK3, KLK13 and PRSS27). Interestingly, Fibrinogen alpha chain (FGA) and Prekallikrein (KLKB1), both exclusively secreted by liver cells, also showed a high correlation in urine. Several clusters consisted of proteins located in the same subcellular compartments, such as lysosomes (PRCP, GSUB, GGH, CPQ, NHLRC3 and IFI30, FSTL1) and azurophil granules in neutrophils (MPO, AZU1). Examples for known molecular interactions were observed, for instance, by PDZK1 and SCLC9A3R1, which are known to interact in proximal tubule cells^23^, and by IST1 and MITD1, both binding to the endosomal sorting complex III in multiple cell types^24,25^.

**Figure 3:**
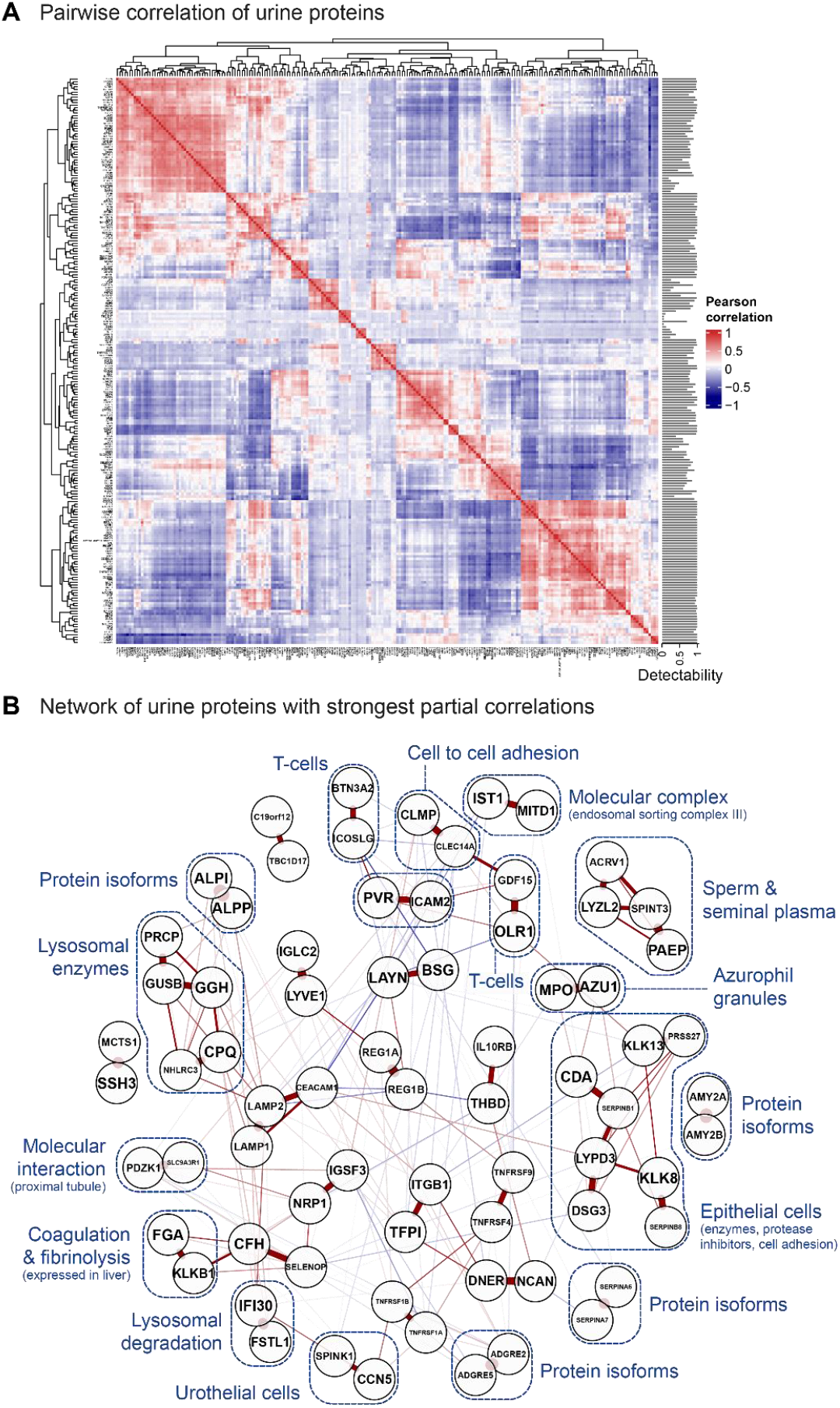
Correlation networks reveal co-regulation of urine proteins. **(A)** Pearson correlation matrix of dilution-corrected urine protein levels. Only proteins with at least one pairwise correlation of r > 0.7 are shown (n=217). Bars on the right indicate detectability for each protein. **(B)** Regularized partial correlation network of urine proteins, annotated with biological mechanisms underlying protein co-regulation. Edge colors indicate the direction of partial correlations (red: positive; blue: negative), and edge thickness represents correlation strength.

## Discussion

Our study shows that the Olink Explore 3072 platform, originally developed for plasma proteomics, can successfully be used in urine and enables comprehensive characterization of protein detectability, variability, and associations. Despite being optimized for circulating proteins, nearly one-quarter of the library showed high detectability (>80%) in urine, with strong concordance between independent batches and good overlap with mass spectrometry data. These highly detectable proteins were enriched for extracellular, exosomal, and cell-surface localization, in line with previous MS-based studies^26–28^. Cell type enrichment identified contributions from hepatocytes, immune cells, and proximal tubule cells, reflecting the potential of urine as a readout for both kidney-specific and systemic processes^29^. Sex was the largest contributor to urinary protein variability, with nearly 1,000 associated proteins reflecting anatomical and physiological differences: prostate-derived proteins (KLK3, MSMB) in men and proteins from vaginal epithelium (CRNN) and hormonal changes (FSHB) in women, the latter capturing systemic hormonal changes comparable to plasma^8^. The correlation network revealed biologically meaningful protein clusters corresponding to shared cellular compartments, tissue origins, and known molecular interactions. We identified four strongly correlated protein isoform pairs, some with evidence for biological co-regulation (e.g., co-expression in immune cells for ADGRE5/ADGRE2^30^), while for others, antibody cross-reactivity due to high sequence homology cannot be ruled out, which is a known potential limitation of affinity-based methods^31^. Although the Olink platform targets preselected proteins and may thereby miss biomarkers not represented in its library, we observed strong concordance with mass spectrometry studies, suggesting that the panel captures a representative and relevant subset of the urine proteome.

Overall, we conclude that the Olink technology is well-suited for urine proteomics with large sample sizes and yields biologically plausible results. Our comprehensive characterization provides detailed information on urine proteins” detectability, variability, and demographic associations to serve as a resource for future targeted studies of the urine proteome.

## Supporting information

Supplementary Material (excl. tables)

## Data Availability

Comprehensive summary data are provided in the supplementary materials, including: protein de-tectability values in urine overall and by batch, as well as stratified by sex (Supplementary Tables 3 and 10), protein variability measures (Supplementary Table 6), associations with demographic fac-tors (Supplementary Table 8), and correlation/network analyses (Supplementary Table 9). Data underlying the results and figures presented in this manuscript are available in the supplementary tables. Code used for statistical analyses is available from the corresponding author upon reasona-ble request. Information on publicly available external datasets used in this study is provided in the Methods and Supplementary Methods sections. Individual-level data from the GCKD study can be made available to approved collaborators upon request to the GCKD data access committee, in ac-cordance with German data protection laws and participant consent agreements. Details are availa-ble at https://www.gckd.org/.

## Acknowledgements

The work of M.M., S.H., O.B., and A.K. was funded by German Research Foundation (DFG) project ID 431984000 (SFB 1453). Germany”s Excellence Strategy (CIBSS, EXC-2189, project ID 390939984) supported the work of Y.L. and A.K.

The GCKD study was and is supported by the BMBF (FKZ 01ER 0804, 01ER 0818, 01ER 0819, 01ER 0820 and 01ER 0821) and the KfH Foundation for Preventive Medicine. Unregistered grants to support the study were provided by corporate sponsors (listed at https://gckd.org). We are grateful for the willingness of the patients to participate in the GCKD study. The enormous effort of the study personnel of the various regional centers is highly appreciated. We thank the large number of nephrologists who provide routine care for the patients and collaborate with the GCKD study. The GCKD investigators are listed in the Supplementary Note.

## Data Sharing Statement

Comprehensive summary data are provided in the supplementary materials, including: protein detectability values in urine overall and by batch, as well as stratified by sex (**Supplementary Tables 3 and 10**), protein variability measures (**Supplementary Table 6**), associations with demographic factors (**Supplementary Table 8**), and correlation/network analyses (**Supplementary Table 9**). Data underlying the results and figures presented in this manuscript are available in the supplementary tables. Code used for statistical analyses is available from the corresponding author upon reasonable request. Information on publicly available external datasets used in this study is provided in the Methods and Supplementary Methods sections. Individual-level data from the GCKD study can be made available to approved collaborators upon request to the GCKD data access committee, in accordance with German data protection laws and participant consent agreements. Details are available at https://www.gckd.org/.

## Author Contributions

Design of this study: S.H., A.K.

Recruitment and management of study: K.U-E., A.K.

Bioinformatics and statistical analysis: M.M., Y.L., O.B., B.U., S.H.

Interpretation of Results: M.M., S.H., A.K.

Wrote the manuscript: M.M., M.S., S.H.

Critically read and approved the manuscript: M.M., M.S., Y.L., O.B., B.U., P.S., C.W., H.M., K.-U.E, O.S., A.K., S.H.

## Generative AI language models

During the preparation of this work, the authors used generative AI language models (Claude Sonnet 4.5, Anthropic) to improve the readability and language of the manuscript. After using this tool, the authors reviewed and edited the content as needed and take full responsibility for the content of the publication.

