## Supplementary Material (excl. tables) for "Characterization of the Urine Proteome Using the Olink Explore 3072 Platform"

| | |
|---|---|
| Anna Köttgen, MD MPH<br>Institute of Epidemiology and Prevention<br>Medical Center – University of Freiburg<br>Hugstetter Str. 49, 79106 Freiburg, Germany<br> | Stefan Haug, MD<br>Institute of Epidemiology and Prevention<br>Medical Center – University of Freiburg<br>Hugstetter Str. 49, 79106 Freiburg, Germany<br> |

### Table of Contents

*Supplementary Methods* ______ *3*

Study design and participants ______ 3

Proteomic measurement, processing and quality control ______ 3

Comparison to mass spectrometry proteomics from urine ______ 4

Protein variability ______ 4

Enrichment analyses ______ 4

Correlation analyses ______ 5

*Supplementary Results* ______ *7*

Sex specific protein detectabilities ______ 7

*Supplementary Figures* ______ *8*

Supplementary Figure 1: Detectability of Olink Explore 3072 proteins in urine by sub-panel ______ 8

Supplementary Figure 2: Correlation of protein measurements across Olink sub-panels ______ 9

Supplementary Figure 3: GO-Term enrichment for highly detectable proteins ______ 8

Supplementary Figure 4: Gene Ontology enrichment for highly variable urine proteins ______ 11

Supplementary Figure 5: Association of urine proteins with BMI and age ______ 12

Supplementary Figure 6: Regularized partial correlation matrix for urine proteins ______ 13

Supplementary Figure 7: Detectability of Olink proteins in urine by sex ______ 14

*References for supplementary material* ______ *17*

### Supplementary Methods

#### Study design and participants

The German Chronic Kidney Disease (GCKD) study is an ongoing prospective observational cohort study that recruited 5,217 adult participants with CKD between 2010 and 2012 across nine German centers[1]. Inclusion criteria were: (1) eGFR between 30-60 ml/min/1.73m² or (2) eGFR >60 ml/min/1.73m² with UACR >300 mg/g or urinary protein-creatinine ratio >500 mg/g. The study was registered in the German Clinical Trials Register (DRKS 00003971) and approved by local ethics committees of all participating institutions (Universities and Medical Faculties of Aachen, Berlin, Erlangen, Freiburg, Hannover, Heidelberg, Jena, München, and Würzburg). All participants provided written informed consent. Biomaterials including DNA and urine samples were collected at baseline and shipped frozen to a central biobank. Detailed descriptions of the study design, standard operating procedures, and participant characteristics have been published previously[1,2].

For the current proteomic analysis, we selected all 1,274 participants with baseline eGFR 45-60 ml/min/1.73m² and UACR <300 mg/g, representing individuals with mildly to moderately decreased glomerular filtration rate and normal to moderately increased albuminuria.

#### Proteomic measurement, processing and quality control

**Sample preparation**: All urine samples were centrifuged at 1,500g for 10 minutes within 30 minutes of donation following standardized GCKD protocols. This centrifugation step removes cells while retaining soluble proteins and exosomes in the supernatant. Supernatants were immediately frozen at -80°C in 9 ml tubes, then thawed once for aliquoting into 100 µl vials for long-term storage. Frozen aliquots were shipped to two independent service facilities: Olink Analysis Service (Uppsala, Sweden) for batch 1 (n=774), and Life and Brain Analysis Service (Bonn, Germany) for batch 2 (n=500).

**Olink Proximity Extension Assay**: Proteins were quantified using the Olink Explore 3072 library with Proximity Extension Assay (PEA) technology[3]. In this antibody-based method, pairs of oligonucleotide-labeled antibodies bind to each target protein. Upon successful dual binding, the oligonucleotide probes come into proximity, hybridize, and form a unique DNA template that is amplified and quantified using next-generation sequencing. Protein abundance is reported as Normalized Protein eXpression (NPX) values on a log2 scale. The Explore 3072 library targets 2,926 unique proteins organized into eight panels: Inflammation I & II, Oncology I & II, Cardiometabolic I & II, and Neurology I & II.

**Optimization for urine**: Prior to the main study, we conducted a pilot experiment using dilution series from 16 urine samples to optimize PEA performance in urine. Testing dilutions of 1:0.1, 1:1, and 1:10, we found that undiluted samples (1:1) achieved optimal protein detection across all panels. Therefore, all study samples were analyzed without dilution.

**Quality control and data processing**: Raw NPX values underwent multi-step quality control. First, we removed 58 proteins (2% of the library) for which 100% of measurements received Olink QC warnings, leaving 2,868 proteins. Six proteins (IL-6, IL-8/CXCL8, TNF, IDO1, LMOD1, and SCRIB) were intentionally measured by four assays each as part of Olink's quality assessment system; these were retained as separate analytes, yielding 2,886 total assays.

**Urine dilution correction**: To account for inter-individual variation in urine concentration, we applied the probabilistic quotient normalization method[4] using highly detectable proteins (detectability >80%) as a stable reference set. Dilution correction was performed separately for all samples included in batch 1 and batch 2. The resulting set of proteins used to compute sample-specific dilution correction factors (pq-factors) included 593 proteins for batch 1 and 659 proteins for batch 2.

**Sample-level quality control**: After dilution correction, we performed principal component analysis on proteins with <20% of values below the limit of detection. Five samples were excluded as multivariate outliers i.e. >5 standard deviations from the mean along any of the first 10 principal components. One additional sample was removed due to excessive QC warnings of >50% of protein measurements flagged. The final dataset comprised 1,268 samples with 2,886 protein analytes representing 2,868 distinct proteins.

#### Protein detectability

Protein detectability is defined as the percentage of urine samples in which the protein was measured above its respective limit of detection as provided by Olink. A detectability of 80% means a protein was found in 4 out of 5 urine samples.

#### Comparison to mass spectrometry proteomics from urine

We compared Olink protein detectability to a comprehensive collection of nine high-resolution mass-spectrometry (MS)-based urine proteome studies[5]. These studies analyzed either cell-free urine or the exosomal fraction from 3-24 healthy donors of both sexes, collectively identifying 8,021 gene products in urine.

All proteins measured by PEA in our study were stratified into ten detectability bins (0-10%, 10-20%, ..., 90-100%), followed by calculation of the percentage of proteins in each bin that had been previously identified in any of the nine MS studies. Protein names were harmonized across all studies using UniProt IDs. Of the 8,021 proteins from the MS compendium, 49 were excluded due to unmappable UniProt IDs, leaving 7,972 proteins for comparison.

#### Protein variability

To quantify protein variability across urine samples, interquartile ranges (IQRs) were calculated for each protein separately in batch 1 and batch 2 using NPX values passing quality control (**Supplementary Table 6**). Combined IQRs represent the median of the two batch-specific IQR values.

#### Enrichment analyses

**Gene Ontology enrichment**: All Olink proteins were annotated with GO-terms using GMT files from g:Profiler[6].

**Cell type and secretome enrichment**: Proteins were annotated with cell types based on single cell RNA expression data from the Human Protein Atlas[7] (HPA), including all cell types where the encoding transcript was among the top 10% of detected transcripts (normalized transcript expression in nTPM, downloaded from https://www.proteinatlas.org/about/download). Each protein was further annotated with its secretome classification from the HPA database[8], creating combined cell type–secretory status labels. Proteins could be assigned to multiple cell types and secretome classes.

**Tissue and compartment enrichment**: Proteins were annotated with tissues and cellular compartments based on immunohistochemistry (IHC) expression grades from the HPA[9] (≥low: low, medium or high intensity; ≥medium: medium or high intensity; ≥high: high intensity).

**Statistical testing**: If not mentioned otherwise, highly detectable proteins (detectability >80%, n=659) were compared to the remaining proteins (detectability ≤80%, n=2,209) using two-sided Fisher's exact tests. P-values were corrected for multiple testing using the Benjamini-Hochberg method. Terms with odds ratios >1 were classified as overrepresented; those with odds ratios <1 were classified as underrepresented.

#### Correlation analyses

**Data preprocessing**: To focus on biological protein co-regulation independent of technical and demographic confounders, we first regressed out effects of dilution, kidney function, demographics, and batch from all protein measurements. Specifically, for each protein, we fitted a linear model including the dilution correction factor (pq-factor), ln(eGFR), ln(UACR), age, sex, and batch (coded as 0 and 1) as covariates. Residuals from these models were then mean-centered and scaled to unit variance.

**Pairwise correlations**: Pearson correlation coefficients were calculated for all possible protein pairs using the standardized residuals with pairwise-complete observations. To focus on strongly co-regulated proteins, we selected all proteins showing at least one correlation with r>0.7 with any other protein, resulting in 217 proteins. These proteins were subjected to unsupervised hierarchical clustering using complete linkage and visualized as a correlation heatmap[10].

##### Regularized partial correlation network

To distinguish direct from indirect protein-protein relationships and reduce false-positive correlations, we estimated regularized partial correlations using the graphical Least Absolute Shrinkage and Selection Operator (LASSO) approach[11]. This method estimates a sparse inverse covariance (precision) matrix, where non-zero elements represent conditional dependencies between proteins after controlling for all other proteins in the network. Graphical LASSO requires a complete data matrix and missing values in the 217 selected proteins were imputed using K-nearest neighbors (KNN) imputation as implemented in the impute.knn function from the R package "impute" (version 1.68.0) with default settings (k=10 neighbors, Euclidean metric).

We then used the estimateNetwork function from the R package “bootnet”[11] (version 1.6) with the following parameters:

- Regularization method: EBICglasso (graphical LASSO with Extended Bayesian Information Criterion model selection)
- Tuning parameter: $\gamma = 5$ (favoring a very sparse, high-specificity network)

- Base correlation: Pearson correlation
- Sample size estimation: pairwise-average

The EBIC-based regularization automatically selects an optimal sparsity level by penalizing model complexity, and the tuning parameter $\gamma = 5$ represents a conservative choice that favors network specificity over sensitivity. The LASSO penalty sets many partial correlations to exactly zero during estimation, reducing false-positive associations. The complete regularized partial correlation matrix for all 217 proteins is visualized as a heatmap in **Supplementary Figure 6**, and all values are provided in **Supplementary Table 9**.

To focus on the strongest direct associations, we selected proteins with at least one pairwise partial correlation >0.3 (n=72) and visualized their partial correlation network using the qgraph package with a modified Fruchterman-Reingold algorithm for node placement (**Figure 3B**). We systematically annotated clusters and protein pairs in the network by screening for: (i) known protein-protein interactions (STRING database[12]), (ii) shared cellular compartments and molecular functions (GeneCards[13]), and (iii) co-expression in specific cell types or tissues (Human Protein Atlas[7–9], proteinatlas.org). These biological annotations are indicated in **Figure 3B**.

### Supplementary Results

#### Sex-specific protein detectabilities

Examining sex-specific detectabilities (**Supplementary Table 10)**, we found a nearly identical detectability distribution for men and women, with a median detectability of 24% for both (**Supplementary Figure 7**). Overall, 254 proteins show a detectability difference of over 10% between men and women; while a significant association with sex can be present, this is not always indicated by a detectability difference. For example, KLK3 and MSMB, which are highly significantly associated with male sex have similar detectabilities in men and women whereas FSHB (significantly associated with female sex) shows a 35% higher detectability in women.

### Supplementary Figures

#### Supplementary Figure 1: Detectability of Olink Explore 3072 proteins in urine by sub-panel

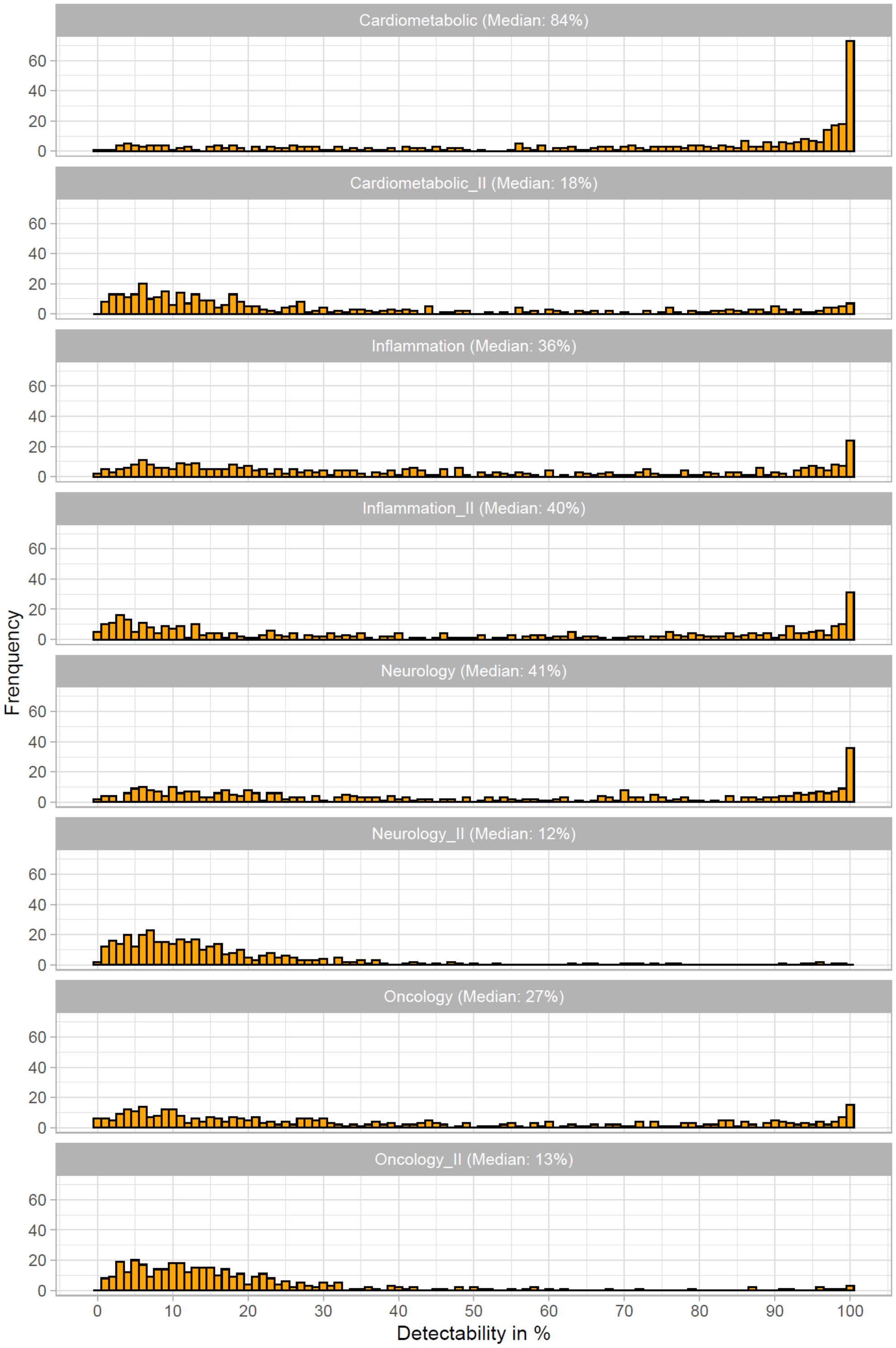


Distribution of protein detectability stratified by the eight Olink Explore 3072 sub-panels: Cardiometabolic I & II, Inflammation I & II, Neurology I & II, and Oncology I & II. Median detectability for each sub-panel is shown in the panel headers.

**Supplementary Figure 2: Correlation of protein measurements across Olink sub-panels**

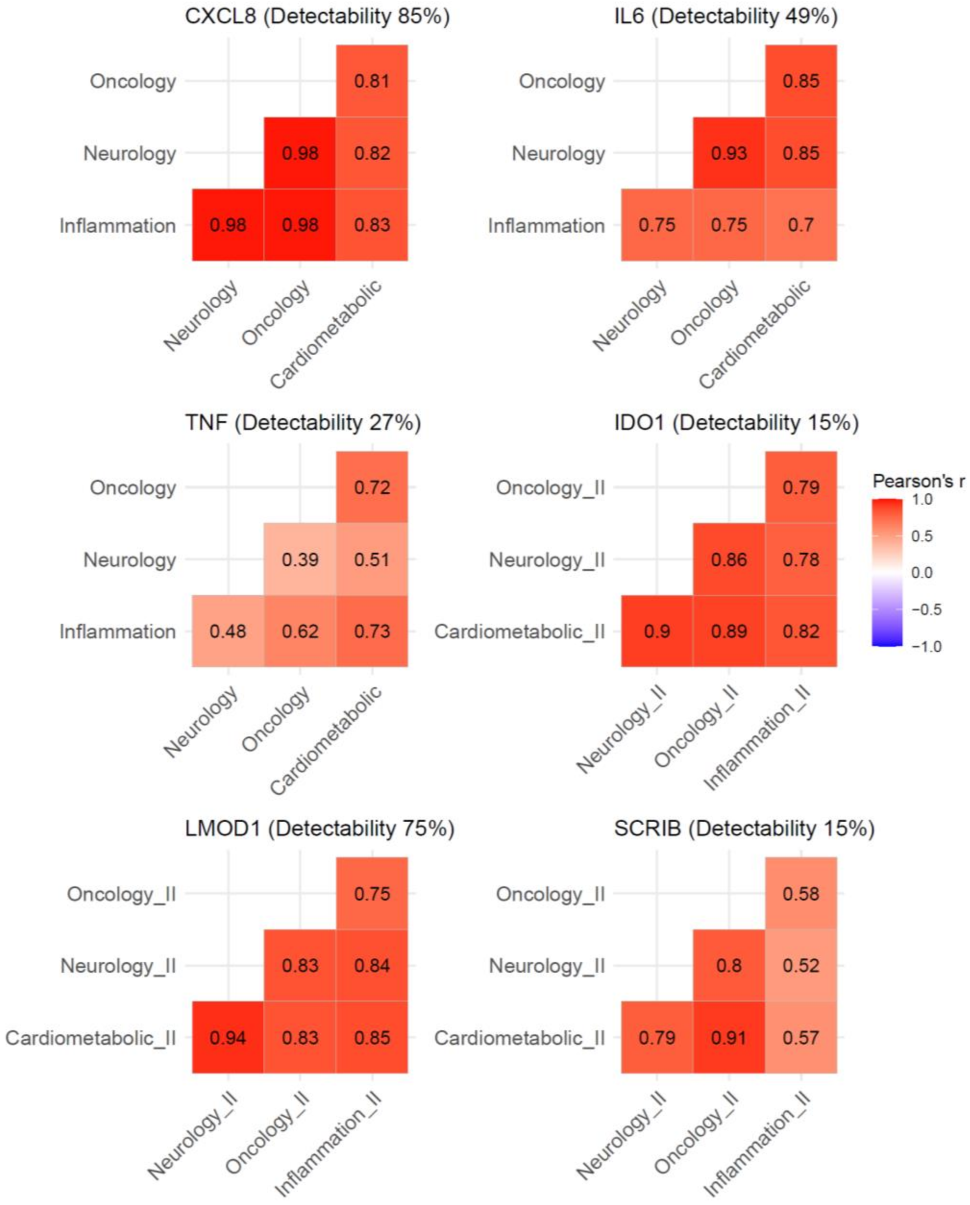


Six proteins (CXCL8, IL-6, TNF, IDO1, LMOD1, and SCRIB) were quantified by four independent assays, each belonging to a different Olink sub-panel. The plots show pairwise Pearson correlations between measurements from different sub-panels for each protein. Overall detectability (percentage above limit of detection) was calculated across all four assays for each protein. Correlations are based on raw Normalized Protein Expression (NPX) values, including measurements below the limit of detection.

**Supplementary Figure 3: GO-Term Enrichment for highly detectable proteins**

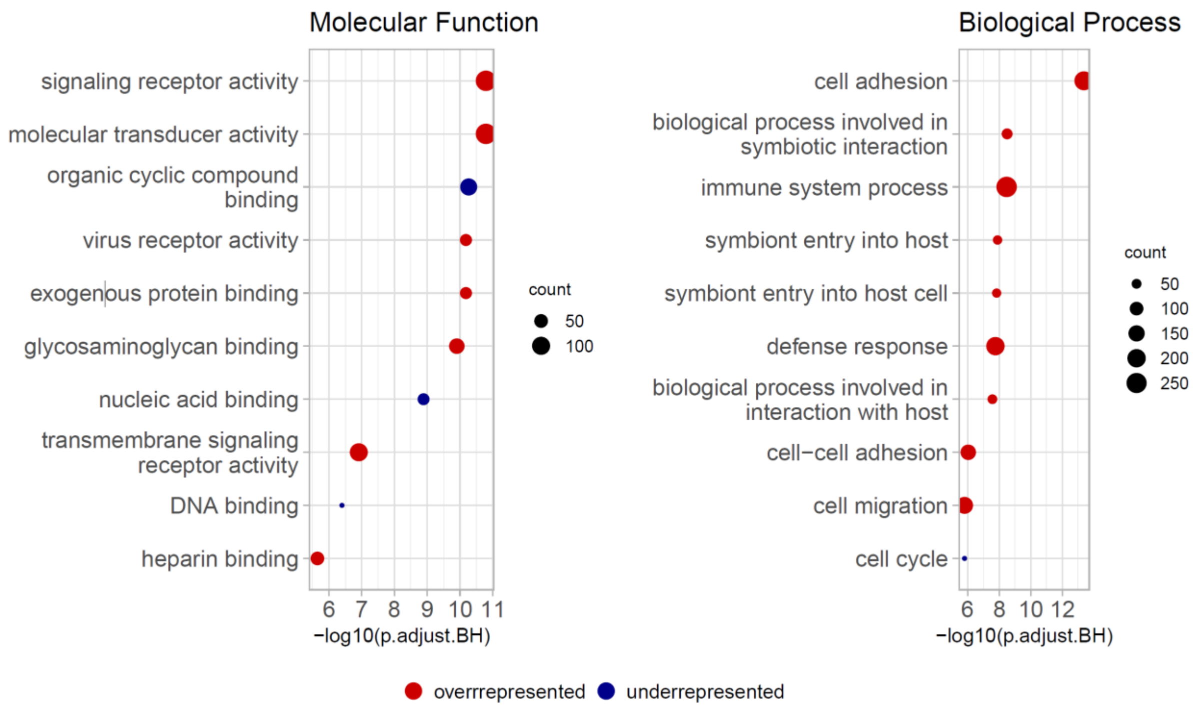


GO-Term Enrichment of molecular functions and biological processes among highly detectable proteins.

#### Supplementary Figure 4: Gene Ontology Enrichment for highly variable urine proteins

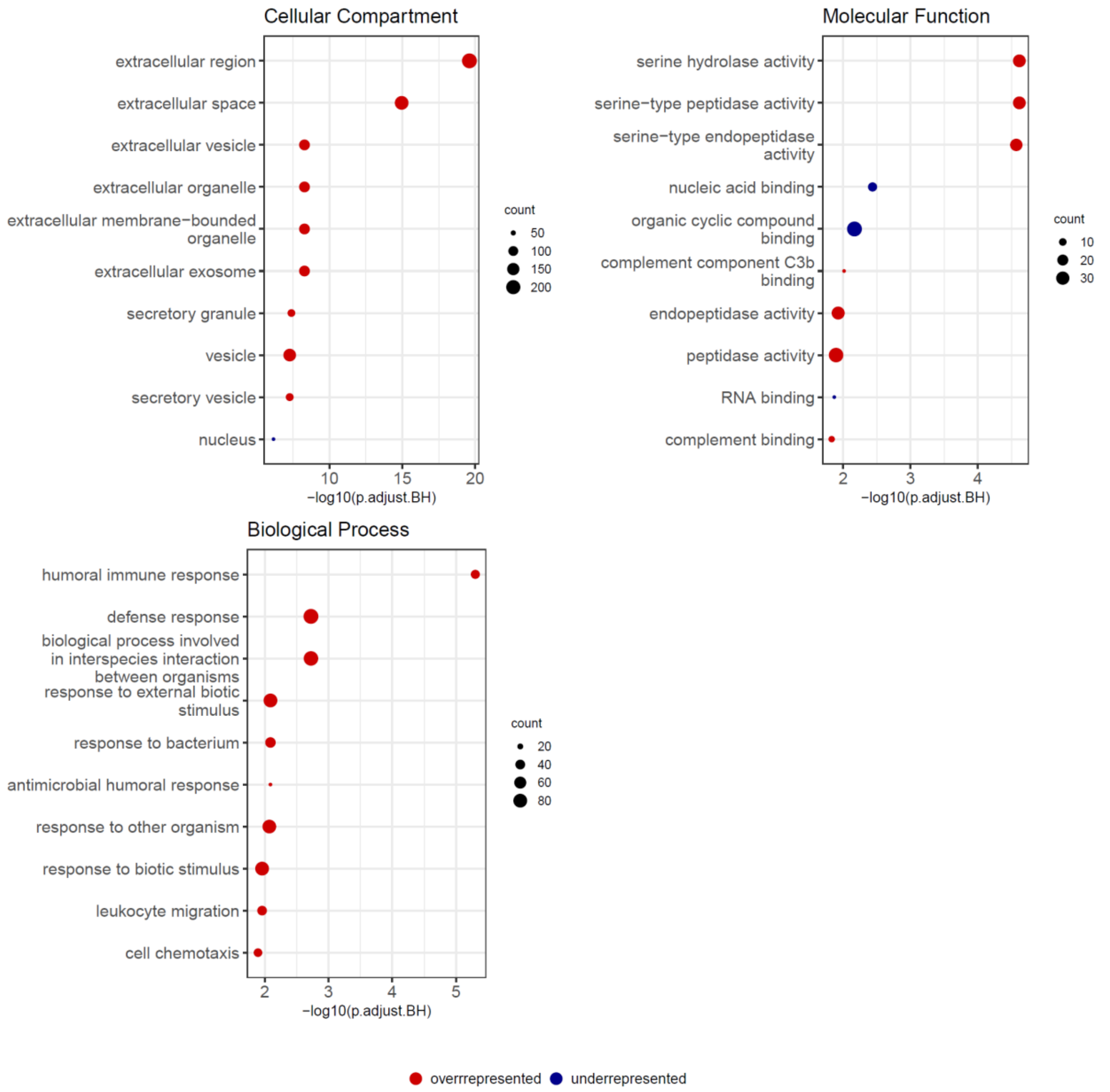


Gene Ontology (GO) Term enrichment analysis for cellular compartments, molecular functions, and biological processes among proteins with high inter-individual variability. Proteins were ranked by interquartile range (IQR) of dilution-corrected NPX values (combined IQRs from batch 1 and batch 2; Supplementary Methods), and the top 10% most variable proteins were compared to the remaining 90%.

#### Supplementary Figure 5: Association of urine proteins with BMI and age

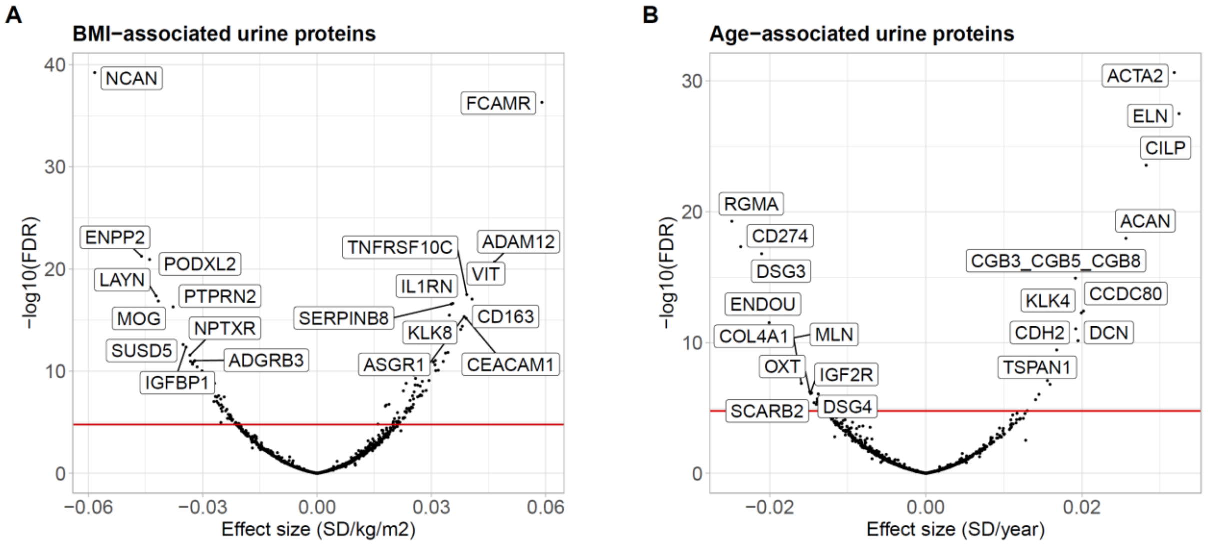


**(A)** Association of dilution-corrected urine protein levels with BMI from multiple linear regression models adjusted for age, sex, eGFR, UACR, and batch. The horizontal red line indicates the significance threshold (FDR = 0.05). Effect sizes are shown as standardized coefficients (SD, standard deviation units). **(B)** Association of dilution-corrected urine protein levels with age from multiple linear regression models adjusted for sex, BMI, eGFR, UACR, and batch. Significance threshold and effect size representation as in panel A. Elastin (ELN) levels in urine show a strong positive association with age, reflecting the same association pattern from blood plasma[14,15]. Plasma elastin levels correlate with various aging indicators in humans and elastin derived fragments were reported to have life-span shortening effects in mice[14].

**Supplementary Figure 6: Regularized partial correlation matrix for urine proteins**

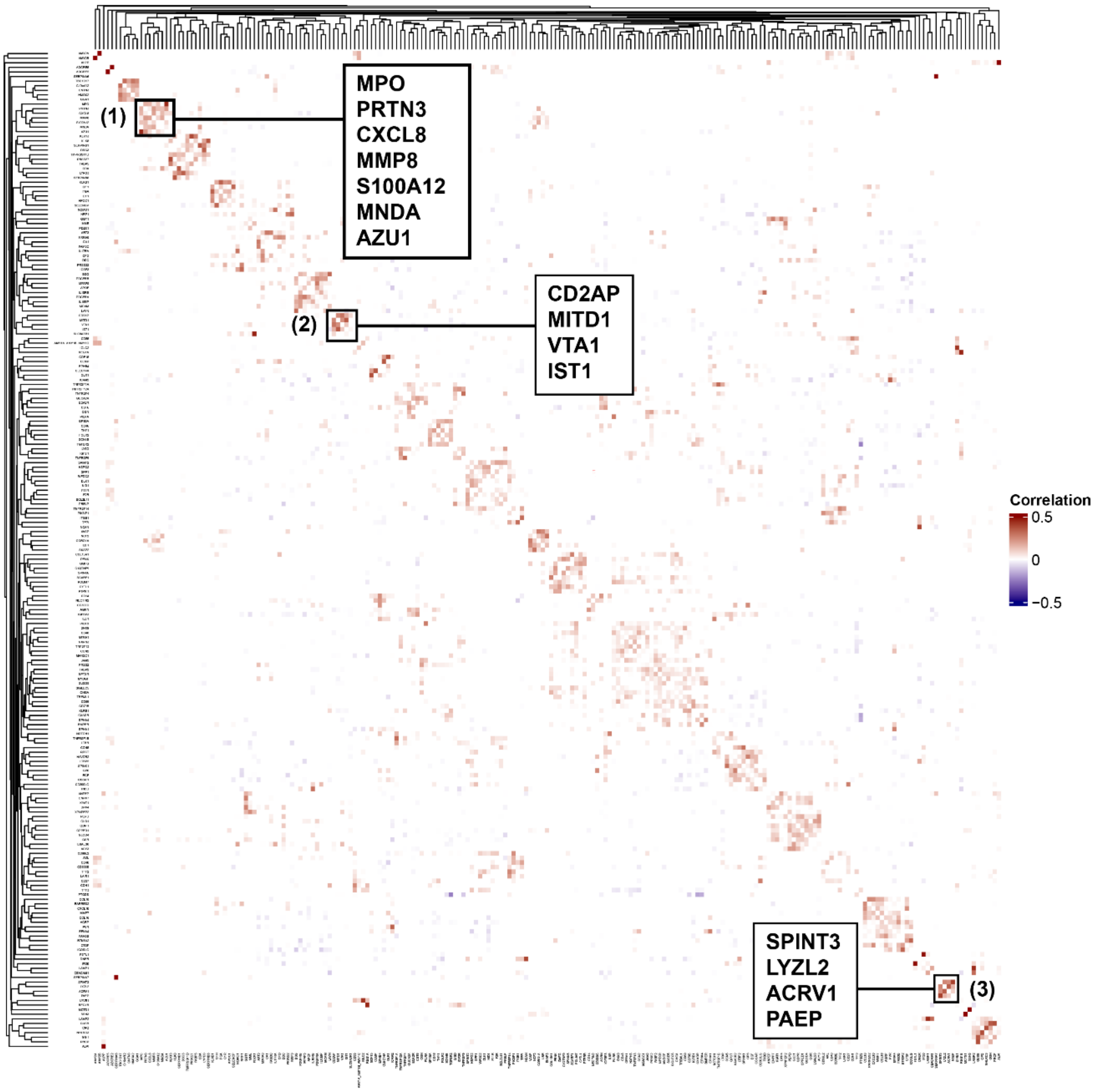


Heatmap of regularized partial correlations between urine proteins, estimated using the EBICglasso approach (Supplementary Methods). The analysis included proteins with at least one Pearson correlation of r > 0.7 with another protein (n=217), based on residuals adjusted for dilution, eGFR, UACR, age, and sex. Proteins were ordered by unsupervised hierarchical clustering using complete linkage. Partial correlation values are provided in **Supplementary Table 9**.

Three black boxes highlight prominent clusters including the indicated proteins. **Cluster (1)** contains correlated proteins expressed in granulocytic cells (S100A12), involved in azurophilic granule degranulation (CXCL8), and proteins found within azurophilic neutrophil granules (MPO, PRTN3, MMP8, MNDA, AZU1). **Cluster (2)** contains proteins (CD2AP, MITD1, VTA1, IST1) related to endosome formation and the endosomal sorting complex III (ESCRT-III), with known molecular interaction between MITD1 and IST1 at ESCRT-III[16]. **Cluster (3)** includes four proteins expressed in the testis (LYZL2, ACRV1), epididymis (SPINT3), and seminal vesicle (PAEP), all potentially co-occurring due to the presence of seminal fluid in urine samples from some male participants.

**Supplementary Figure 7: Detectability of Olink proteins in urine by sex**

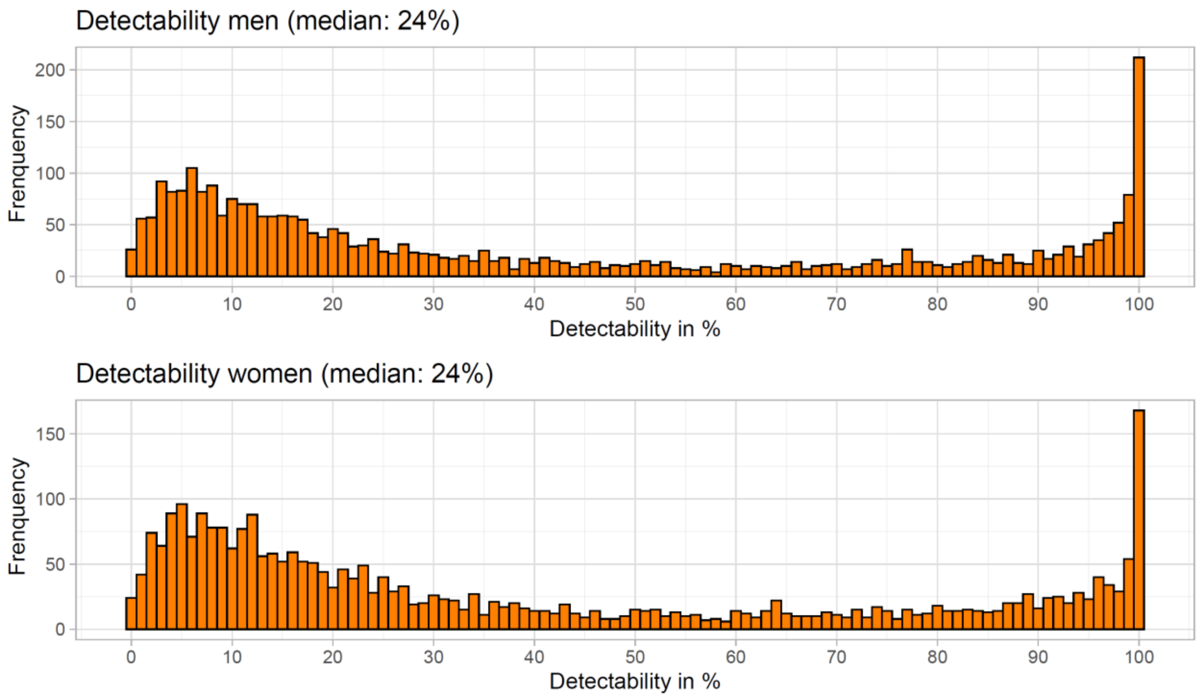


Distribution of protein detectability stratified by sex.

### Supplementary Acknowledgements

#### List of GCKD Study Investigators

A list of nephrologists currently collaborating with the GCKD study is available at http://www.gckd.org.

| | |
|---|---|
| University of Erlangen-Nürnberg | Kai-Uwe Eckardt, Heike Meiselbach, Markus Schneider, Mario Schiffer, Hans-Ulrich Prokosch, Barbara Bärthlein, Andreas Beck, André Reis, Arif B. Ekici, Susanne Becker, Ulrike Alberth-Schmidt, Anke Weigel, Sabine Marschall |
| University of Freiburg | Gerd Walz, Anna Köttgen, Ulla Schultheiß, Wibke Bechtel-Walz, Fruzsina Kotsis, Simone Meder, Erna Mitsch, Ursula Reinhard |
| RWTH Aachen University | Jürgen Floege, Rafael Kramann, Turgay Saritas |
| Charité, University Medicine Berlin | Elke Schaeffner, Seema Baid-Agrawal, Kerstin Theisen |
| Hannover Medical School | Kai Schmidt-Ott |
| University of Heidelberg | Martin Zeier, Claudia Sommerer |
| University of Jena | Gunter Wolf, Martin Busch, |
| Ludwig-Maximilians University of München | Thomas Sitter |
| University of Würzburg | Christoph Wanner, Vera Krane, Britta Bauer |
| Medical University of Innsbruck, Division of Genetic Epidemiology | Florian Kronenberg, Julia Raschenberger, Barbara Kollerits, Lukas Forer, Sebastian Schönherr, Hansi Weissensteiner |
| University of Regensburg, Institute of Functional Genomics | Peter Oefner, Wolfram Gronwald |
| Department of Medical Biometry, Informatics and Epidemiology (IMBIE), University of Bonn | Matthias Schmid, Jennifer Nadal |
